# Anaplastic Large Cell Lymphoma (ALCL) and hernia mesh implants: an unrecognised association

**DOI:** 10.1101/2025.06.12.25329183

**Authors:** Andrew Ross, Emily R. James, Indrani Karpha, Elizabeth Ryan, Anita J. M. Kalakonda, Igor Racu-Amoasii, Nagabhushan Seshadri, Julia Salmeron-Villalobos, Itziar Salaverria, Arvind Arumainathan, Gladstone A. Amos Burke, Joseph R. Slupsky, Suzanne D. Turner, Nagesh Kalakonda

## Abstract

The risk of ALK-ve ALCL associated with textured breast implants (BIA-ALCL) is a notable example of lymphomas associated with medical devices. We describe 2 cases of ALK-ve ALCL associated with meshes, commonly used devices in surgical procedures such as herniorrhaphy. Our cases, each presenting some 10 years following mesh insertions, show anatomical and causal association to index tumours. In addition, we show that the genetic profile has significant overlaps with BIA-ALCL, invoking similar etiopathogenesis. Hence, we propose that mesh implant associated-ALCL (MIA-ALCL) is an unrecognised association and unreported disease entity that merits recognition, broader investigation, and surveillance.

## Background

Since an anecdotal case report of anaplastic large cell lymphoma (ALCL) presenting in close proximity to a breast implant in 1997(Keech and Creech 1997) further reports followed(Clemens, *et al* 2024, Miranda, *et al* 2019). Two decades elapsed before acknowledgement of breast implant associated (BIA)-ALCL as a standalone entity within the revised World Health Organisation (WHO) classification of haematological malignancies(Swerdlow, *et al* 2016). BIA-ALCL is a clear public health concern and since 2011 has led to management guidelines(Clemens, *et al* 2019, Expert Panel on Breast, *et al* 2023, Mehta-Shah, *et al* 2018, Turton, *et al* 2021), medical device alerts from national regulatory agencies such as the FDA (US) and MHRA (UK)(De Jong, *et al* 2021), and withdrawal of some implants from the market. Whilst the etiopathogenetic mechanisms of BIA-ALCL are yet to be fully established, it is clear that the common denominator in all cases is a textured breast implant(De Jong, *et al* 2021, Turner, *et al* 2020). Chronic inflammation is postulated to play a key role as with other medical device related malignancies.

We report two cases of anaplastic lymphoma kinase (ALK) negative ALCL (ALK-ve ALCL), who were both 70 years of age, associated with hernia meshes presenting at a single centre. The clinical presentations, radiological, histopathological, and genetic features bear a significant resemblance to BIA-ALCL and invoke a causal association with mesh implants. We submit that the data presented attests to the existence of an as yet unreported entity, i.e., mesh implant associated ALCL (MIA-ALCL), that merits appreciation and wider investigation.

### Case reports

**Patient 1** presented in 2014 with an ulcerated mass in his left inguinal region. Medical history included inguinal herniorrhaphy in 2005 (Bard^®^ prolene mesh; batch/lot no. unobtainable), and co-morbidities including hypertension, bronchiectasis, hypercholesterolaemia, and diet-controlled Type 2 diabetes mellitus. A magnetic resonance (MR) scan revealed a 6×6×2.5 cm nodal mass in direct contact with the mesh (**Figure1; patient 1; A and B**). Histopathology and immunohistochemistry (IHC) of the biopsy revealed ‘hallmark’ tumour cells consistent with a diagnosis of ALK-ve ALCL (**Figure 2**). Tumour embedded ‘criss-cross’ mesh fibres were clearly visible in the haematoxylin and eosin (H&E)-stained histology section (**Figure 2A)**. The malignant cells were positive for CD45, CD30 and BCL2; with patchy expression of c-myc (~40%), MUM1, BCL6 and Granzyme B; but negative for ALK1, PAX5, TIA-1, EBV-LMP1 and EBER, CD2, CD3, CD5, CD10, CD15, CD19, CD20, CD56, CD57, CD79a, CD138, CCND1, MNF116, S100, Melan A, SOX10 and HMB45. The Ki-67 proliferation index was 40% and the ulcerated lesion was MRSA +ve. Additionally, investigations revealed hepatitis B core antibody seropositivity but no active infection. A baseline ^18^fluoro-deoxy-glucose positron-emission and computed tomography (^*18*^*FDG-PET-CT)* scan was consistent with stage IIA disease with intensely FDG avid left inguinal and external iliac nodes (SUV_max_ 39.3). Despite 2 cycles of CHOP chemotherapy (cyclophosphamide, hydroxydaunorubicin, vincristine and prednisolone), parenteral antibiotics and topical wound care there was evidence of local progression (**Figure 1 C-F**), ongoing weight loss, and deterioration of ECOG performance status (necessitating parenteral feeding). To achieve disease control, the patient was referred for involved field radiotherapy (18Gy; 3 fractions) and subsequently commenced brentuximab vedotin (BV) monotherapy (enrolled into, and consents obtained, as part of the post-authorisation safety (BV) ARROVEN study (PASS: MA25101)).

**Figure 1.**
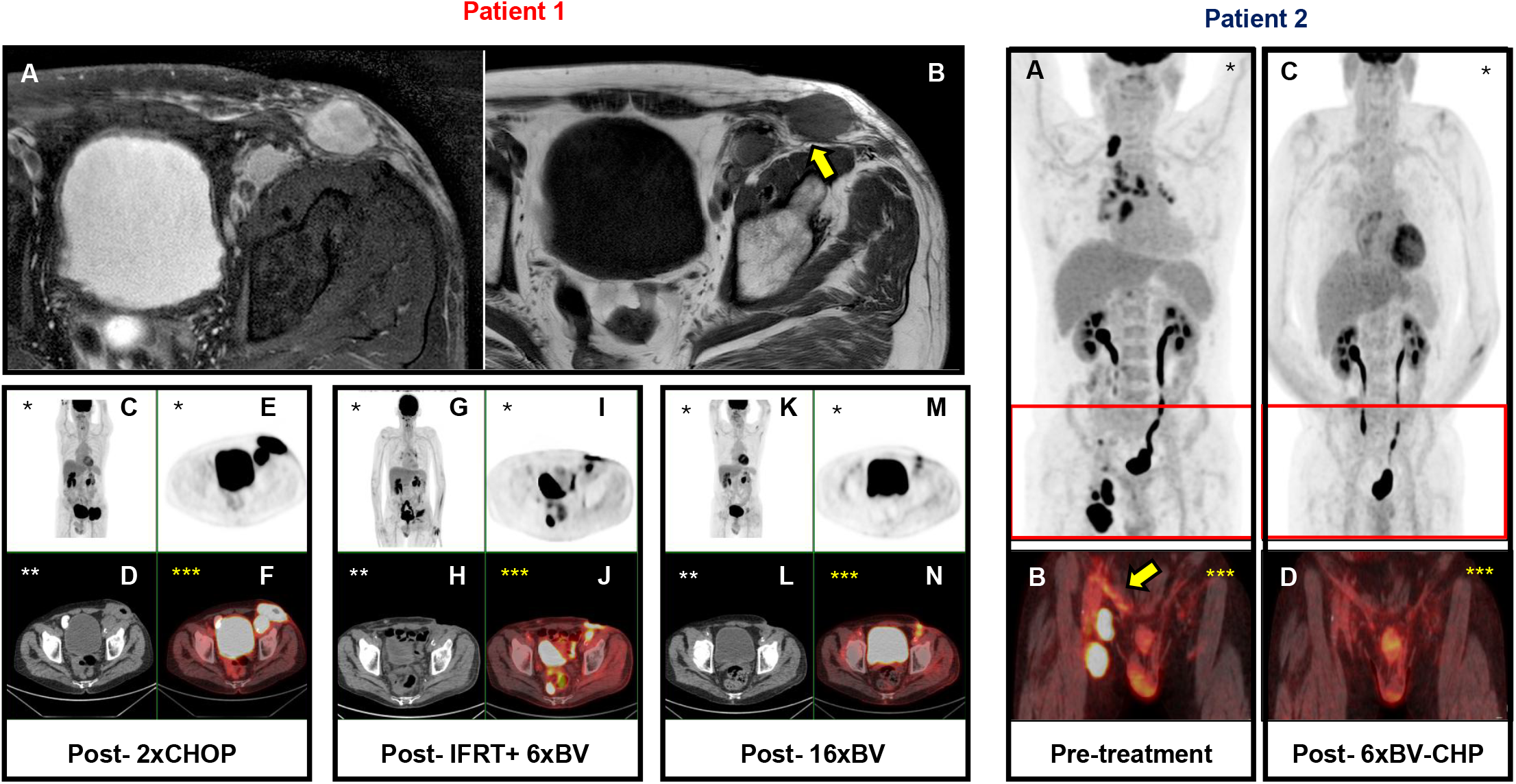
MRI and ^18^FDG-PET/CT scans reveal intertwined inflamed mesh and tumour. Patient 1: **A:** Pre-treatment axial T2-weighted MR image shows the inguinal nodes. **B:** The mesh abutting and underlying the mass (arrow) is highlighted in the T1-weighted image. **C – N:** Whole body MIP, axial PET/-CT, and fused PET-CT images of the pelvis at defined treatment stages as indicated below the panels. **Patient 2: A:** Pre-treatment whole body PET-CT MIP image showing stage III disease with prominent right groin and mediastinal nodes. **B:** The arrow in the fused PET-CT coronal slice highlights the inflamed mesh. **C, D**: Corresponding post-treatment MIP and fused PET-CT images confirm a complete metabolic response. *(FDG-Fluorodeoxyglucose, MIP-Maximum Intensity Projection, BV-Brentuximab vedotin, C-cyclophosphamide, H-hydroxydaunorubicin, O-oncovin, P-prednisolone, IFRT-involved field radiotherapy; * PET alone, **CT alone, *** fused PET + CT)*.

**Figure 2.**
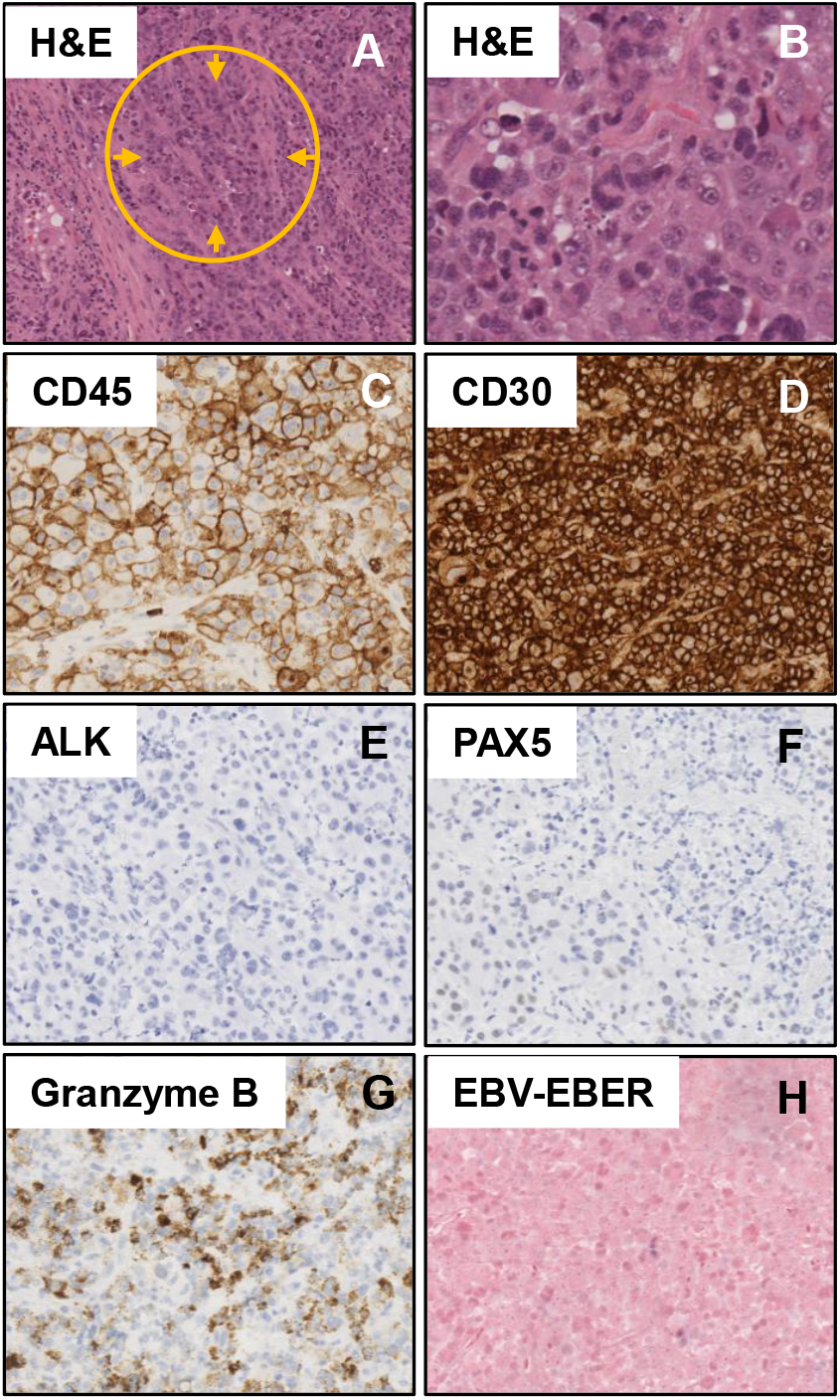
Histopathological characterization of an inguinal node biopsy (from patient 1) reveals tumour cells intertwined with mesh material. **A:** Haematoxylin and eosin (H&E) staining of tissue. The circle and arrows highlight the ‘crisscross’ mesh fibres embedded by tumour cells and **B:** Higher magnification shows the neoplastic cells including large hallmark cells with horseshoe-shaped nuclei. **C – H:** Immunohistochemical staining with the indicated antibodies showing CD45 and CD30 expression on tumour cells but a lack of ALK, PAX5, and EBV-EBER staining, and patchy Granzyme-B uptake. *(Magnifications: A – 10x; B-H – 40x)*.

Tumour regression was apparent after 2 cycles of BV and the healing, but open, wound now revealed a protruding mesh. The mesh overhang was surgically excised but removal in its entirety was deemed impossible due to fibrosis and tissue assimilation. An FDG PET-CT scan post 6x BV showed good partial remission (**Figure 1 G-J**). The end of treatment PET-CT (post 16 cycles) showed excellent metabolic response (**Figure 1 K-N**). The residual low-grade FDG uptake was considered to be due to ongoing inflammation of the mesh/healing scar. The patient maintained a stable complete remission but in 2020 unfortunately succumbed to COVID19.

**Patient 2** presented during the COVID19 pandemic (2020) with a rapid-onset (2-week history) right upper thigh mass on a background of predominantly cardiac co-morbidities (9 years prior; Medtronic (Covidien) composite parietex™ mesh; lot no. SLG00200). Regular medications included warfarin, furosemide, atorvastatin, cyanocobalamin, salbutamol and steroid inhalers. An MRI scan revealed a 4.5×3×4 cm right groin nodal mass. Biopsy showed monomorphous, medium-sized lymphoid cells with occasional ‘hallmark’ cells expressing CD45, CD30 and MUM1 but negative for ALK1, CD20, PAX5, TdT, CCND1, CD10, BCL6 and EBV. Patchy expression of CD3, CD4 and CD5 was thought to be due to a reactive T-cell infiltrate. The features were consistent with ALK-ve ALCL (Ki-67 90% +ve). A baseline FDG PET-CT scan **(Figure 1, Patient 2, A)** revealed Stage III disease (supraclavicular, mediastinal, portocaval, and right inguinal nodes). The *in-situ* right sided hernia mesh showed moderate FDG uptake consistent with inflammation (**Figure 1; patient 2; A and B**). The patient was treated with 6 cycles of B-CHP (BV, cyclophosphamide, attenuated doxorubicin (75%), and prednisolone) without dose delays or reductions. Following an excellent response noted on an interval CT, the end-of treatment PET-CT scan (**Figure 1, patient 2; C and D**) confirmed complete metabolic remission which has been maintained to date.

### WES and FISH analyses

Archived formalin fixed and paraffin embedded (FFPE) biopsy material from patient 1 (human tissue ethics approval no. 07-Q0104-16, UK) was obtained for whole exome sequencing (WES) analyses (see **methods and Figure S1 in supplementary data**). Additional Fluorescence *in situ* hybridisation (FISH) analysis was conducted (details of genes and probes in supplementary **Table S1**) using established protocols (see **supplementary methods**). The tissue from patient 1 was only sufficient for one sequencing run. As mitigation, and to account for lack of genomic DNA for comparison, WES performed had a median coverage of over 200x with 90% of loci covered by over 100 reads (supplementary **Figure S1A**). The biopsy from patient 2 was friable and failed to yield sufficient or useful material for genomic analyses. Despite patient consent, a second biopsy in this patient was not feasible due to constraints imposed by the COVID pandemic and urgency to initiate treatment.

The WES mutational profile from patient 1 with MIA-ALCL shows significant overlap with genes mutated in BIA-ALCL, cutaneous and systemic ALK-ve ALCL **(Figure 3)**. For clarity, the reported gene mutations/alterations indicated for sALCL, cALCL and BIA-ALCL in **Figure 3A** are a composite representation from the literature and not from single cases. WES analysis revealed impactful *JAK1* (truncating; R174*; results in JAK1/STAT3 pathway activation) and *TP53* (missense; H193Y; leads to loss of p53 function) mutations with Variable Allele Frequencies (VAF): 0.507 and 0.365 respectively) (**Figures 3A and B**).

**Figure 3.**
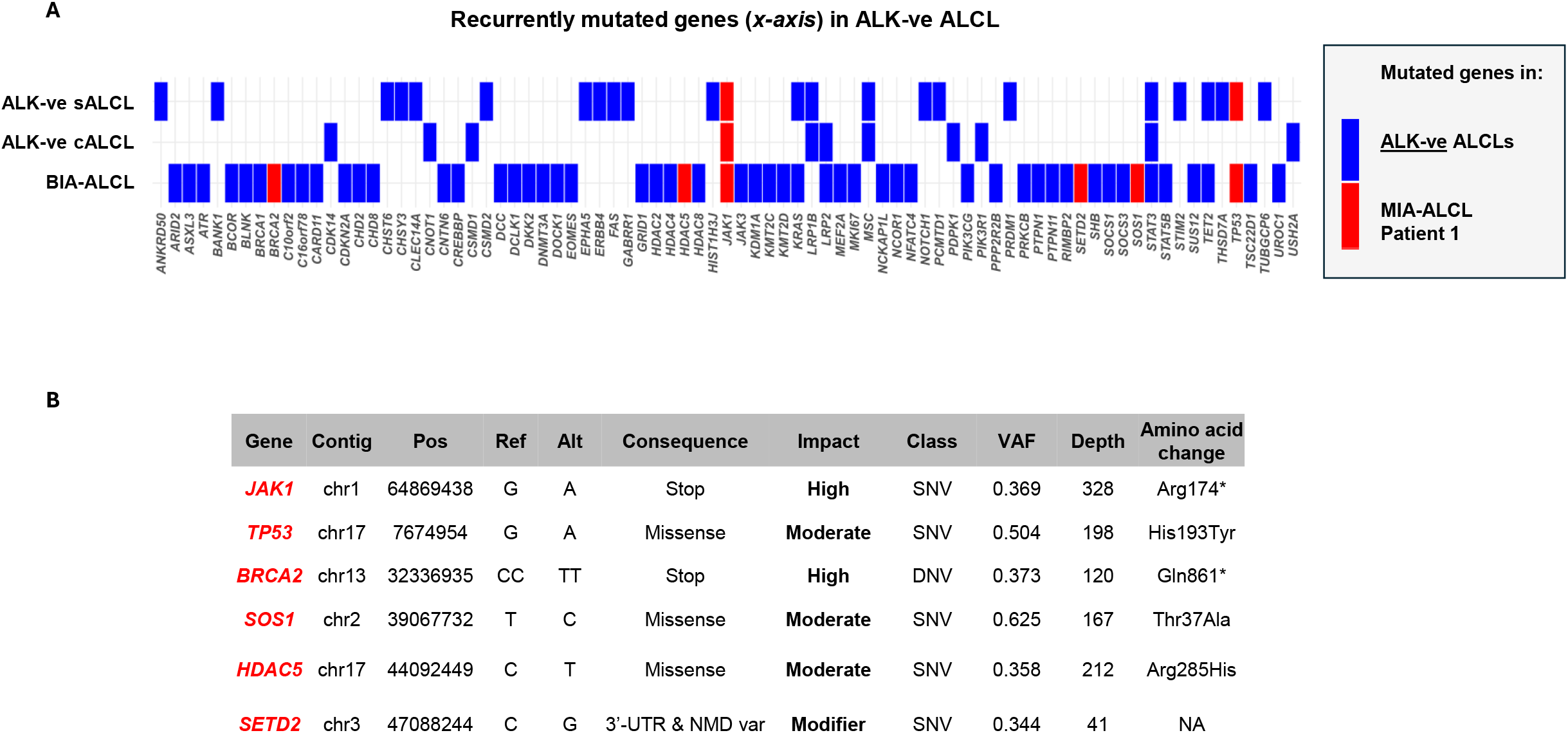
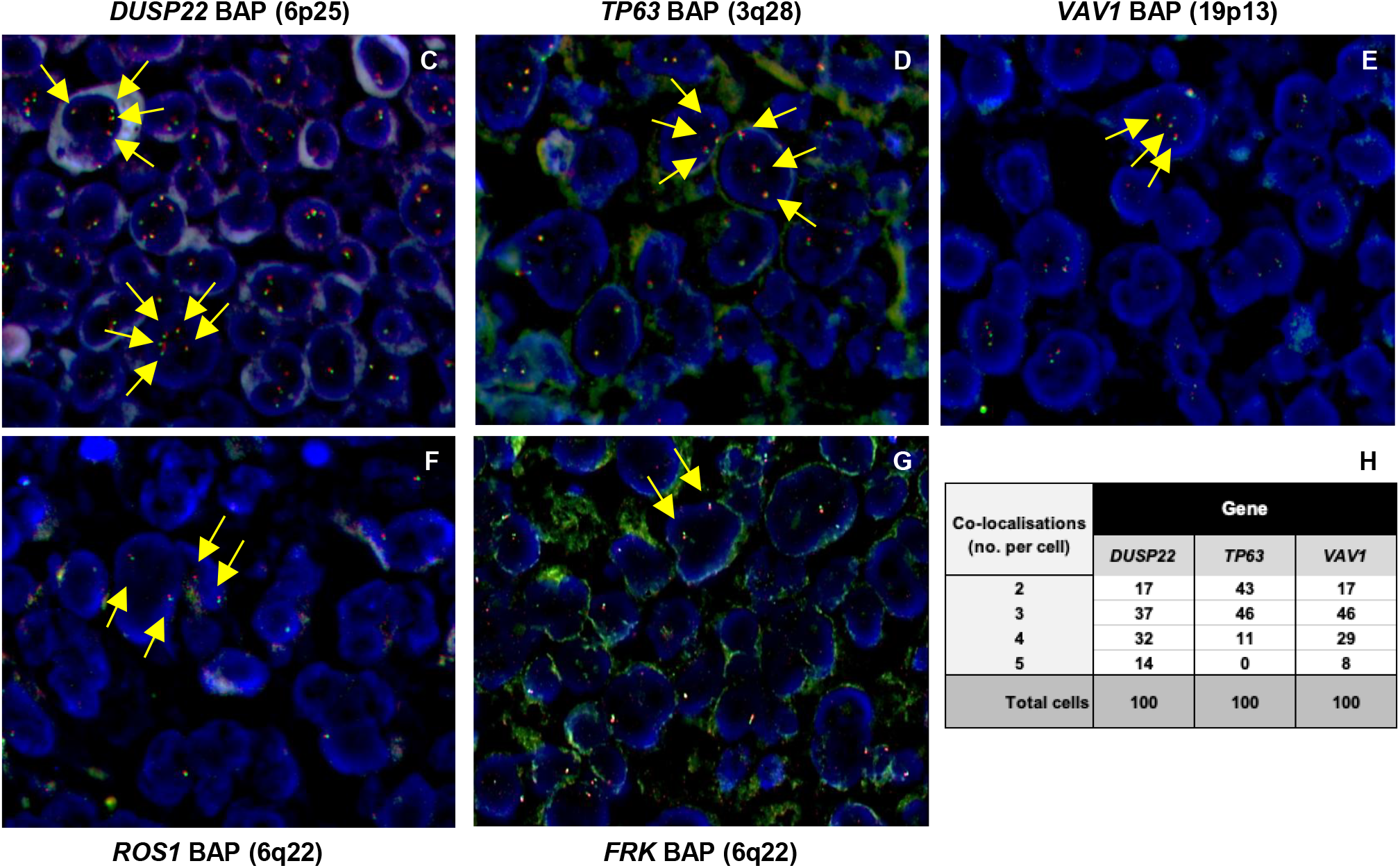
Whole Exome Sequencing (WES) and Fluorescence *in situ* hybridization (FISH) analysis of biopsy tissue from patient 1. **A:** Comparison of reported (**composite data from multiple patients**) mutations within indicated genes in 3 subtypes of ALK-ve ALCL (Breast implant associated (BIA), cutaneous (c) and systemic/nodal (s) ALK-ve ALCL) shown in **blue**. The mutations found in patient 1 with MIA-ALCL are shown in **red**. The accompanying table **(B)** shows the gene mutations found in patient 1 with indicated nucleotide changes, consequence, impact, variant allele frequency (VAF) and amino acid changes (*SNV – single nucleotide variant, DNV – double nucleotide variant, UTR – untranslated region, NMD – nonsense mediated decay*). **C-G:** extra colocalizations (marked with yellow arrows) of *DUSP22, TP63*, and *VAV1* loci; but two normal colocalizations of *ROS1* and *FRK* loci. **H**. The table shows the number of cells with multiple copies (2-5) of *DUSP22, TP63* and *VAV1* genes (100 cells were counted for each gene).

Additional mutations of *BRCA2, SOS1, HDAC5* and *SETD2* were also seen (**Figure 3B**). A list of other variants detected is presented in **supplementary Table S2**. FISH analyses (**Figure 3; C-G**) of a panel of 5 genes was performed, based on insights from ALK-ve ALCL(Feldman, *et al* 2023), did not reveal any chromosomal translocations but showed extra colocalizations corresponding to extra copies of *DUSP22* (6p25), *TP63* (3q28) and *VAV1* (19p13) (**Figure 3H** shows percentage of cells with 2 – 5 CNGs) but only two copies of *ROS1* and *FRK* genes, both located at 6q22. The results indicate significant overlap with genomic abnormalities seen in BIA-ALCL.

## Discussion

The cases of MIA-ALCL presented above show significant similarities to BIA-ALCL and mirror its clinical presentation, histopathological features, disease course and response to treatment(Horwitz, *et al* 2019, Mehta-Shah, *et al* 2018, Turton, *et al* 2021). Lag times in our patients (~10 years since herniorrhaphy) and variable proliferative rates (40 and 90%) again are akin to reports of BIA-ALCL. It is striking that the lymphoma subtype in both patients is also ALK-ve ALCL (as with BIA-ALCL). In both cases, the adjacency and proximity of meshes to index lesions, and features of chronic inflammation and/or infection are noteworthy. Hence, in both cases the hernia meshes are clearly a common underlying factor for the emergence of ALK-ve ALCL. Of note, genomic aberrations in patient 1 (especially *JAK1* and *TP53* mutations, and extra copies of *DUSP22, TP63* and *VAV1*) overlap with genes and pathways implicated in BIA-ALCL(Akkad, *et al* 2023, Laurent, *et al* 2020, Los-de Vries, *et al* 2020, Quesada, *et al* 2021). It has previously been shown in BIA-ALCL that an inflammatory microenvironment, polyclonal invasion of Th17/Th1 cells subsets, inflammatory cytokines and JAK-STAT3 pathway mutations likely lead to malignant transformation. The data presented suggests that similar factors may be at play in MIA-ALCL(Fitzal, *et al* 2019, Turner 2019, Turner, *et al* 2020). Of significance, the occurrence of MIA-ALCL is not restricted to a single type of mesh or manufacturer.

BIA-ALCL has garnered significant media and professional attention, but such interest was not established or of consequence for over a decade since early reports. Besides breast implants, other medical devices (including orthopaedic prostheses, vascular grafts, valve replacements, dental implants, silicone injections) have also been linked to both lymphoid and non-lymphoid malignancies including ALCL, diffuse large B-cell lymphoma (DLBCL) and squamous cell carcinoma (SCC)(Di Napoli, *et al* 2024, Santanelli di Pompeo, *et al* 2024). However, although the mechanisms responsible are yet to be fully understood, potential factors include chronic inflammation and/or infections (including those arising from biofilms and the microbiome) and implant characteristics (texture, composition, microparticle shedding, etc)(Foppiani, *et al* 2023, Turner, *et al* 2020). The potential for genetic predisposition, especially in the case of BIA-ALCL, is also an area of active research(Deva, *et al* 2020).

Early generation Bard^®^ and Medtronic™ meshes have been widely used for hernia-repairs for decades. ‘Monofilament” meshes are microporous (pore sizes of 0.8mm) and designed to stimulate fibrosis and tissue integration(Baylon, *et al* 2017). Although effective in reinforcing a weakened abdominal/inguinal wall, the mesh can be a nidus for chronic inflammation/infections. Whether some facet of the meshes themselves and/or chronic inflammation is responsible for MIA-ALCL remains to be determined. Despite the emergence of newer generations, including those purported to be bio-compatible, cost implications likely preclude widespread adoption to reduce associated risks.

Our enquiries to date, of similar occurrences in larger patient cohorts (e.g., the UK Chemo-T trial(Gleeson, *et al* 2018) and within other institutions), have not yet identified additional cases mostly due to a lack of annotations of a history of herniorrhaphy and/or paucity of patients presenting with early-stage disease where an association may be more clear-cut.

Unlike BIA-ALCL where an association should be immediately obvious, a history of herniorrhaphy may not be sought, forthcoming nor volunteered, especially in the event of a significant lag time. In addition, a causal association may not be appreciated if the disease presents at an advanced stage. As a case in point, patient 2 presented with a rapid onset, two-week history (likely due to the high proliferation rate (Ki67 90%), with infra-and supradiaphragmatic (stage III) nodes. But for the ‘inflamed’ mesh noted in the PET/CT scan, the association would be easily missed. As with BIA-ALCL where treatment outcomes are excellent, BV-based treatment in our MIA-ALCL cases induced stable remissions.

The importance of completely excising the mesh, as is recommended in the treatment of BIA-ALCL (*en bloc* capsulectomy removing both the implant and the fibrous capsule), as both a primary treatment and to prevent relapse is unknown(Clemens, *et al* 2018). Whereas breast implants can be surgically extricated, removal of hernia meshes is nigh impossible as they are designed to be ‘self-fixing’ and induce ‘tissue tethering’ and due to potential sequelae.

In conclusion, we propose that MIA-ALCL is a distinct clinical entity that requires the attention of oncologists, surgeons, healthcare practitioners, mesh manufacturers and regulatory bodies. We suggest that a broader investigation, retrospective and prospective surveillance and research efforts are warranted.

## Data Availability

Reasonable requests for sequencing and FISH and any other data should be directed to the corresponding author.

## Author contributions

SDT, GAAB, JRS, and NK: Study conceptualisation. ERJ: WES, genomic analysis, literature review. AR, IK, ER, AK, AA and NK: patient consents, samples/image/case reviews, patient care and literature reviews. AA: local investigator for ARROVEN study. IS, JS: FISH studies, analysis and interpretation of the results. I R-A: Histopathology. NS: Radiology input. All authors have contributed to the writing/editing of the manuscript and approve its contents.

## Acknowledgements

We are grateful to the two patients for their consent and involvement. SDT is supported by research funding from a CRUK Cambridge Centre [C9685/A25117] award, a European Union Horizon 2020 Marie Skłodowska-Curie Doctoral Training Network grant (no. 101072735) and an EU National Institute for Cancer Research Programme (EXCELES, no. LX22NPO5102). ERJ is supported by a UK-MRC doctoral training award. AK was supported by funding from a NIHR Clinical Research Fellowship. AR and IK are funded by Clatterbridge Research Doctoral Fellowships. NK and JS receive funding support from Merseyside against blood cancers charity (MABC, The Bloom appeal, Registered Charity no. 1157459). We thank Dr Kathryn Scott (University of Liverpool, UK) for critical reading of the manuscript.

## Supplementary data

Supplementary methods, figures (S1, S2), and tables (S1, S2)

## Data statement and availability

Reasonable requests for WES, FISH and any other data should be directed to the corresponding author. These potential adverse events have been reported to the MHRA, UK

## References

Akkad, N., Kodgule, R., Duncavage, E.J., Mehta-Shah, N., Spencer, D.H., Watkins, M., Shirai, C. & Myckatyn, T.M. (2023) Evaluation of Breast Implant-Associated Anaplastic Large Cell Lymphoma With Whole Exome and Genome Sequencing. Aesthet Surg J, 43, 318–328.

Baylon, K., Rodriguez-Camarillo, P., Elias-Zuniga, A., Diaz-Elizondo, J.A., Gilkerson, R. & Lozano, K. (2017) Past, Present and Future of Surgical Meshes: A Review. Membranes (Basel), 7.

Clemens, M.W., Brody, G.S., Mahabir, R.C. & Miranda, R.N. (2018) How to Diagnose and Treat Breast Implant-Associated Anaplastic Large Cell Lymphoma. Plast Reconstr Surg, 141, 586e–599e.

Clemens, M.W., Jacobsen, E.D. & Horwitz, S.M. (2019) 2019 NCCN Consensus Guidelines on the Diagnosis and Treatment of Breast Implant-Associated Anaplastic Large Cell Lymphoma (BIA-ALCL). Aesthet Surg J, 39, S3–S13.

Clemens, M.W., Myckatyn, T.M., Di Napoli, A., Feldman, A.L., Jaffe, E.S., Haymaker, C.L., Horwitz, S.M., Hunt, K.K., Kadin, M.E., McCarthy, C.M., Miranda, R.N., Prince, H.M., Santanelli di Pompeo, F., Holmes, S.D. & Phillips, L.G. (2024) American Association of Plastic Surgeons Consensus on Breast Implant-Associated Anaplastic Large-Cell Lymphoma. Plast Reconstr Surg, 154, 473–483.

De Jong, W.H., Panagiotakos, D., Proykova, A., Samaras, T., Clemens, M.W., De Jong, D., Hopper, I., Rakhorst, H.A., Santanelli di Pompeo, F., Turner, S.D., S.E.a. & Scheer (2021) Final opinion on the safety of breast implants in relation to anaplastic large cell lymphoma: Report of the scientific committee on health, emerging and environmental risks (SCHEER). Regul Toxicol Pharmacol, 125, 104982.

Deva, A.K., Turner, S.D., Kadin, M.E., Magnusson, M.R., Prince, H.M., Miranda, R.N., Inghirami, G.G. & Adams, W.P., Jr. (2020) Etiology of Breast Implant-Associated Anaplastic Large Cell Lymphoma (BIA-ALCL): Current Directions in Research. Cancers (Basel), 12.

Di Napoli, A., Fruscione, S., Mazzola, S., Amodio, R., Graziano, G., Mannino, R., Zarcone, M., Bertolazzi, G., Bonaccorso, N., Sciortino, M., De Bella, D.D., Savatteri, A., Belluzzo, M., Norrito, C.A., Sparacino, R., Contiero, P., Tagliabue, G., Costantino, C. & Mazzucco, W. (2024) Emerging Non-Breast Implant-Associated Lymphomas: A Systematic Review. Cancers (Basel), 16.

Expert Panel on Breast, I., Chetlen, A., Niell, B.L., Brown, A., Baskies, A.M., Battaglia, T., Chen, A., Jochelson, M.S., Klein, K.A., Malak, S.F., Mehta, T.S., Sinha, I., Tuscano, D.S., Ulaner, G.A. & Slanetz, P.J. (2023) ACR Appropriateness Criteria(R) Breast Implant Evaluation: 2023 Update. J Am Coll Radiol, 20, S329–S350.

Feldman, A.L., Laurent, C., Narbaitz, M., Nakamura, S., Chan, W.C., de Leval, L. & Gaulard, P. (2023) Classification and diagnostic evaluation of nodal T- and NK-cell lymphomas. Virchows Arch, 482, 265–279.

Fitzal, F., Turner, S.D. & Kenner, L. (2019) Is breast implant-associated anaplastic large cell lymphoma a hazard of breast implant surgery? Open Biol, 9, 190006.

Foppiani, J.A., Raska, O., Taritsa, I., Hernandez Alvarez, A., Lee, D., Escobar-Domingo, M.J., Berger, J., Klener, P., Schuster, K.A., Abdo, D., Clemens, M.W. & Lin, S.J. (2023) Incidental Bystander or Essential Culprit: A Systematic Review of Bacterial Significance in the Pathogenesis of Breast Implant-Associated Anaplastic Large Cell Lymphoma. Int J Mol Sci, 25.

Gleeson, M., Peckitt, C., To, Y.M., Edwards, L., Oates, J., Wotherspoon, A., Attygalle, A.D., Zerizer, I., Sharma, B., Chua, S., Begum, R., Chau, I., Johnson, P., Ardeshna, K.M., Hawkes, E.A., Macheta, M.P., Collins, G.P., Radford, J., Forbes, A., Hart, A., Montoto, S., McKay, P., Benstead, K., Morley, N., Kalakonda, N., Hasan, Y., Turner, D. & Cunningham, D. (2018) CHOP versus GEM-P in previously untreated patients with peripheral T-cell lymphoma (CHEMO-T): a phase 2, multicentre, randomised, open-label trial. Lancet Haematol, 5, e190–e200.

Horwitz, S. & O’Connor, O.A. & Pro, B. & Illidge, T. & Fanale, M. & Advani, R. & Bartlett, N.L. & Christensen, J.H. & Morschhauser, F. & Domingo-Domenech, E. & Rossi, G. & Kim, W.S. & Feldman, T. & Lennard, A. & Belada, D. & Illés, Á. & Tobinai, K. & Tsukasaki, K. & Yeh, S.-P. & Shustov, A. & Hüttmann, A. & Savage, K.J. & Yuen, S. & Iyer, S. & Zinzani, P.L. & Hua, Z. & Little, M. & Rao, S. & Woolery, J. & Manley, T. & Trümper, L. & Aboulafia, D. & Advani, R. & Alpdogan, O. & Ando, K. & Arcaini, L. & Baldini, L. & Bellam, N. & Bartlett, N. & Belada, D. & Yehuda, D.B. & Benedetti, F. & Borchman, P. & Bordessoule, D. & Brice, P. & Briones, J. & Caballero, D. & Carella, A.M. & Chang, H. & Cheong, J.W. & Cho, S.-G. & Choi, I. & Choquet, S. & Colita, A. & Congui, A.G. & D’Amore, F. & Dang, N. & Davison, K. & de Guibert, S. & Brown, P.d.N. & Delwail, V. & Demeter, J. & di Raimondo, F. & Do, Y.R. & Domingo, E. & Douvas, M. & Dreyling, M. & Ernst, T. & Fanale, M. & Fay, K. & Feldman, T. & Ferrero, S.F. & Flinn, I.W. & Forero-Torres, A. & Fox, C. & Friedberg, J. & Fukuhara, N. & Garcia-Marco, J. & Cruz, J.G. & Codina, J.G. & Gressin, R. & Grigg, A. & Gurion, R. & Christensen, J.H. & Haioun, C. & Hajek, R. & Hanel, M. & Hatake, K. & Hensen, R. & Horowitz, N. & Horwitz, S. & Huttmann, A. & Illes, A. & Illidge, T. & Ishizawa, K. & Islas-Ohlmayer, M. & Jacobsen, E. & Janakiram, M. & Jurczak, W. & Kaminski, M. & Kato, K. & Kim, W.S. & Kirgner, I. & Iyer, S. & Kuo, C.-Y. & Lazaroiu, M.C. & Du, K.L. & Lee, J.-S. & LeGouill, S. & Lennard, A. & LaRosee, P. & Levi, I. & Link, B. & Maisonneuve, H. & Maruyama, D. & Mayer, J. & McCarty, J. & McKay, P. & Minami, Y. & Mocikova, H. & Morra, E. & Morschhauser, F. & Munoz, J. & Nagai, H. & O’Connor, O. & Opat, S. & Pettengell, R. & Pezzutto, A. & Pfreundschuh, M. & Pluta, A. & Porcu, P. & Pro, B. & Quach, H. & Rambaldi, A. & Renwick, W. & Reyes, R. & Izquierdo, A.R. & Rossi, G. & Ruan, J. & Rusconi, C. & Salles, G. & Santoro, A. & Sarriera, J. & Savage, K. & Shibayama, H. & Shustov, A. & Suh, C. & Sureda, A. & Tanimoto, M. & Taniwaki, M. & Tilly, H. & Tobinai, K. & Trneny, M. & Trumper, L. & Tsukamoto, N. & Tsukasaki, K. & Vitolo, U. & Walewski, J. & Weidmann, E. & Wilhelm, M. & Witzens-Harig, M. & Yacoub, A. & Yamamoto, K. & Yeh, S.-P. & Yoon, S.-S. & Yuen, S. & Yun, H.J. & Zain, J. & Zinzani, P.L. (2019) Brentuximab vedotin with chemotherapy for CD30-positive peripheral T-cell lymphoma (ECHELON-2): a global, double-blind, randomised, phase 3 trial. The Lancet, 393, 229–240.

Keech, J.A., Jr. & Creech, B.J. (1997) Anaplastic T-cell lymphoma in proximity to a saline-filled breast implant. Plast Reconstr Surg, 100, 554–555.

Laurent, C., Nicolae, A., Laurent, C., Le Bras, F., Haioun, C., Fataccioli, V., Amara, N., Adelaide, J., Guille, A., Schiano, J.M., Tesson, B., Traverse-Glehen, A., Chenard, M.P., Mescam, L., Moreau, A., Chassagne-Clement, C., Somja, J., Escudie, F., Andre, M., Martin, N., Lacroix, L., Lemonnier, F., Hamy, A.S., Reyal, F., Bannier, M., Oberic, L., Prade, N., Frenois, F.X., Beldi-Ferchiou, A., Delfau-Larue, M.H., Bouabdallah, R., Birnbaum, D., Brousset, P., Xerri, L. & Gaulard, P. (2020) Gene alterations in epigenetic modifiers and JAK-STAT signaling are frequent in breast implant-associated ALCL. Blood, 135, 360–370.

Los-de Vries, G.T., de Boer, M., van Dijk, E., Stathi, P., Hijmering, N.J., Roemer, M.G.M., Mendeville, M., Miedema, D.M., de Boer, J.P., Rakhorst, H.A., van Leeuwen, F.E., van der Hulst, R., Ylstra, B. & de Jong, D. (2020) Chromosome 20 loss is characteristic of breast implant-associated anaplastic large cell lymphoma. Blood, 136, 2927–2932.

Mehta-Shah, N., Clemens, M.W. & Horwitz, S.M. (2018) How I treat breast implant-associated anaplastic large cell lymphoma. Blood, 132, 1889–1898.

Miranda, R.N., Medeiros, L.J., Ferrufino-Schmidt, M.C., Keech, J.A., Jr., Brody, G.S., de Jong, D., Dogan, A. & Clemens, M.W. (2019) Pioneers of Breast Implant-Associated Anaplastic Large Cell Lymphoma: History from Case Report to Global Recognition. Plast Reconstr Surg, 143, 7S–14S.

Quesada, A.E., Zhang, Y., Ptashkin, R., Ho, C., Horwitz, S., Benayed, R., Dogan, A. & Arcila, M.E. (2021) Next generation sequencing of breast implant-associated anaplastic large cell lymphomas reveals a novel STAT3-JAK2 fusion among other activating genetic alterations within the JAK-STAT pathway. Breast J, 27, 314–321.

Santanelli di Pompeo, F., Firmani, G., Stanzani, E., Clemens, M.W., Panagiotakos, D., Di Napoli, A. & Sorotos, M. (2024) Breast Implants and the Risk of Squamous Cell Carcinoma of the Breast: A Systematic Literature Review and Epidemiologic Study. Aesthet Surg J, 44, 757–768.

Swerdlow, S.H., Campo, E., Pileri, S.A., Harris, N.L., Stein, H., Siebert, R., Advani, R., Ghielmini, M., Salles, G.A., Zelenetz, A.D. & Jaffe, E.S. (2016) The 2016 revision of the World Health Organization classification of lymphoid neoplasms. Blood, 127, 2375–2390.

Turner, S.D. (2019) The Cellular Origins of Breast Implant-Associated Anaplastic Large Cell Lymphoma (BIA-ALCL): Implications for Immunogenesis. Aesthet Surg J, 39, S21–S27.

Turner, S.D., Inghirami, G., Miranda, R.N. & Kadin, M.E. (2020) Cell of Origin and Immunologic Events in the Pathogenesis of Breast Implant-Associated Anaplastic Large-Cell Lymphoma. Am J Pathol, 190, 2–10.

Turton, P., El-Sharkawi, D., Lyburn, I., Sharma, B., Mahalingam, P., Turner, S.D., MacNeill, F., Johnson, L., Hamilton, S., Burton, C. & Mercer, N. (2021) UK Guidelines on the Diagnosis and Treatment of Breast Implant-Associated Anaplastic Large Cell Lymphoma on behalf of the Medicines and Healthcare products Regulatory Agency Plastic, Reconstructive and Aesthetic Surgery Expert Advisory Group. Br J Haematol, 192, 444–458.

