## Supplementary data for "Anaplastic Large Cell Lymphoma (ALCL) and hernia mesh implants: an unrecognised association"

**Supplementary methods:**

**Whole Exome Sequencing (WES)**: DNA was isolated from FFPE material using deparaffinization solution and the ‘All-Prep DNA/RNA FFPE Kit’ (Qiagen). WES libraries were prepared and captured by KAPA HyperExome probes (Roche) and sequenced across multiple lanes using a NovoSeq 6000 (Illumina) platform (Genomics Core Facility at CEITEC, Masaryk University, Brno, Czech Republic).

Quality metrics of raw FastQ files were obtained using FastQC. Trimming of Illumina standard adapters was conducted by TrimGalore. Reads were aligned to the human genome (hg38); NCBI) using the Burrows-Wheeler Alignment (BWA) tool BWA-mem. Bam files from different lanes belonging to the same sample were merged and coordinate-sorted using Samtools. Duplicate reads arising from artefactual processes were marked by Mark Duplicates (GATK) and removed from further analysis. Base Quality Score Recalibration (GATK) was conducted using information from read groups, dinucleotide context, cycle number, and known SNPs to identify and correct for patterns of systematic errors in base quality scores. Variant calling to detect single nucleotide alterations and small insertions and/or deletions was carried out by MuTect2 (GATK), Octopus, Lofreq and VarScan2. For variant calling with MuTect2, the 1000 genomes panel of normals (PON) was used to detect mapping artefacts and a germline resource containing population allele frequencies of common and rare variants was used to provide prior probabilities for germline variants. GATK’s Learn Read Orientation Model and Filter Mutect calls were used to filter out single strand substitution errors suspected to arise from the formalin fixation process and Illumina NovoSeq machines. All filtered variant calls were normalised and annotated for biological context using the ENSEMBL variant effect predictor (Vep). As there was no matched germline, only variants called across all 4 variant callers were considered. Further filtering of variants was conducted using databases of common population single nucleotide polymorphisms such as dbsnp138 and gnomAD.

**Fluorescence *in situ* hybridisation (FISH)** analyses were performed using standard protocols. Breaks at *DUSP22/IRF4*, *VAV1*, *TP63*, *FRK*, *TYK2* and *ROS1* were analyzed using homemade or commercial FISH probes (Metasystems, Altlußheim, Germany) (**Table S1**). FISH slides were visualized using a Nikon Eclipse 50i (Nikon, Shinagawa, Tokyo) and pictures captured using ISIS software (Metasystems, Altlußheim, Germany).

Figure S1

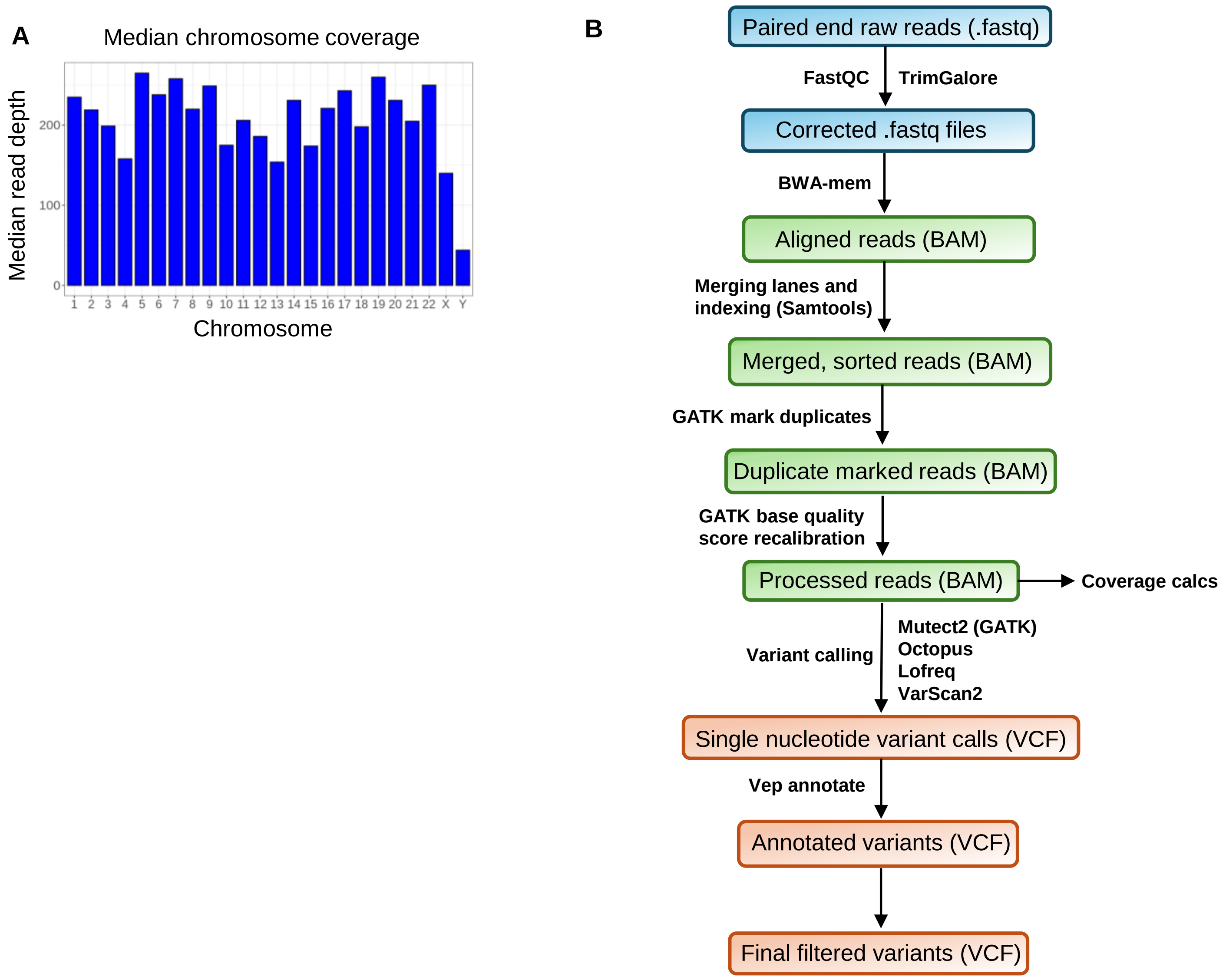


**Figure S1: Whole Exome Sequencing (WES) and analysis**
**A.** Median WES coverage and read depth.
**B.** Bioinformatics analysis pipeline built on Snakemake.

**Figure S2:**

**A**

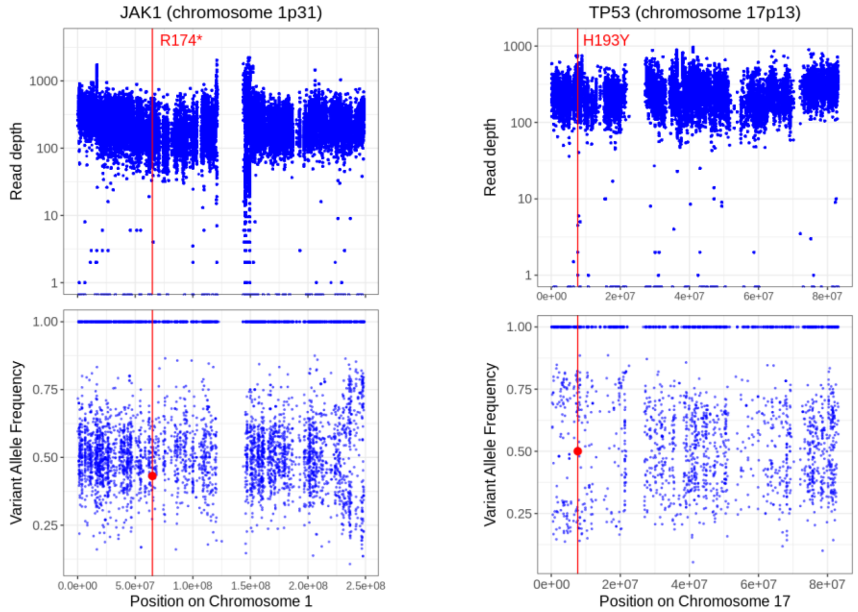


**B**

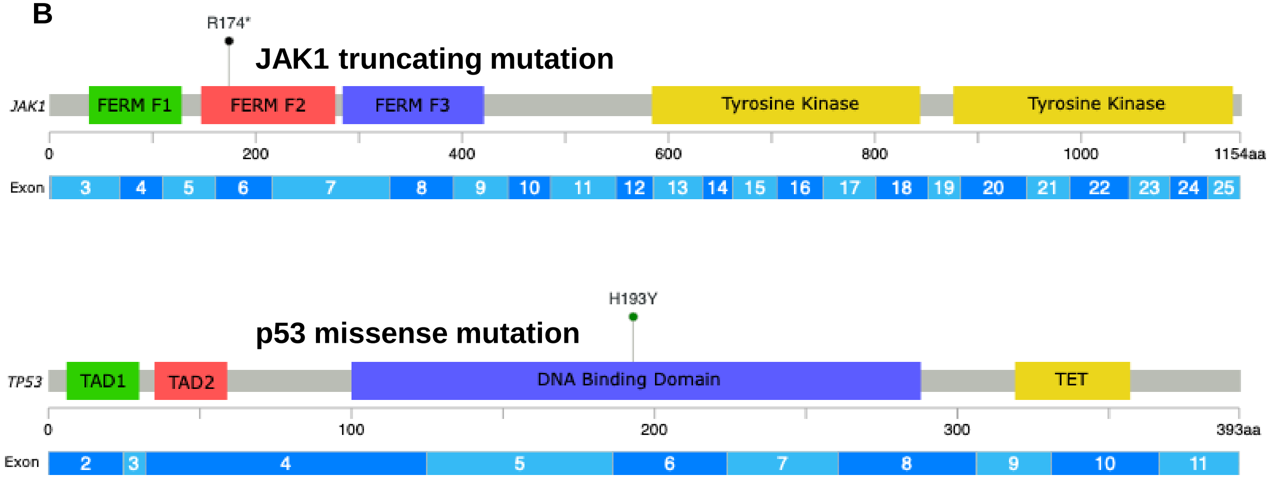


**Figure S2: WES analysis of *JAK1* and *p53* genes**
**A. Read depths (upper) and variant allele frequencies (VAF) (lower)** across chromosome 1 (*left*) and chromosome 17 (*right*). Red lines indicate the position of a R174* (truncation-encoding) mutant in JAK1 (*chr.1p31*) and a missense TP53-H193Y (*chr.17p13*) mutant; red dots indicate average VAF.
**B: Schematic representation of protein-domains and exonic architecture of JAK1 and p53 proteins/genes respectively**. Positions of the mutated amino acids in JAK1 (FERM F2) and p53 (DNA binding) are indicated. *(B – adapted from cBioportal; FERM F1: ubiquitin-like, FERM F2: acyl-CoA binding protein-like, FERM F3: pleckstrin homology-like (canonical tri-lobed FERM domain). TAD1 and 2: transactivating domains, TET: tetramerization motif).*

**Table S1: FISH break apart probes and details**

| Gene | Annotation | Gene Location | BAC 5' | BAC5' location | BAC 3' | BAC 3' LOCATION | Reference |
|---|---|---|---|---|---|---|---|
| ***VAV1*** | hg19 | chr19:6 772,679-6,857,377 | RP11-114A7 | chr19:6 248,889-6,408,433 | RP11-876D1 | chr19:7 211,320-7,445,835 | Adapted from Boddicker et al, Blood 2016 |
| ***DUSP22 (IRF4)*** | hg18 | chr6:237 100-296,353 | RP3-416J7 | chr6:89 702-213,735 | RP5-1077H22 | chr6:835 761-929,128 | Salaverria et al, Blood 2011 |
| ***DUSP22 (IRF4)*** | hg18 | chr6:237 100-296,353 | RP3-416J7 | chr6:89 702-213,735 | RP5-856G1 | chr6:940 429-1,092,223 | Salaverria et al, Blood 2011 |
| ***TP63*** | hg19 | chr3:189 349,216-189,615,068 | CTD-2653J11 | chr3:189 229,868-189,338,362 | CTD-2192L7 | chr3:189 788,306-189,981,208 | Adapted from Vasmatzis et al, Blood 2012 |
| ***TP63*** | hg19 | chr3:189 349,216-189,615,068 | CTD-2653J11 | chr3:189 229,868-189,338,362 | CTD-2513B12 | chr3:189 893,587-190,097,216 | Adapted from Vasmatzis et al, Blood 2012 |
| ***ROS1*** | hg19 | chr6:117 609,530-117,747,018 | XL ROS1-GOPC BA (Metasystems #ref D-6029-100-OG) | | | | Breakpoints described in Crescenzo et al, Cancer Cell 2015 |
| ***FRK*** | hg19 | chr6:116 262,693-116,381,921 | RP11-721G11 | chr6:116 060,027-116,167,934 | RP11-656L18 | chr6:116 405,751-116,560,328 | Adapted from Hu et al, Leukemia 2018 |
| ***FRK*** | hg19 | chr6:116, 262,693-116,381,921 | RP1-188H10 | chr6:116 167,835-116,255,040 | RP11-186J10 | chr6:116 500,585-116,661,498 | Adapted from Hu et al, Leukemia 2018 |


1. Boddicker RL, Razidlo GL, Dasari S, et al. Integrated mate-pair and RNA sequencing identifies novel, targetable gene fusions in peripheral T-cell lymphoma. Blood 2016;128(9):1234-45.
2. Salaverria I, Philipp C, Oschlies I, et al. Translocations activating IRF4 identify a subtype of germinal center-derived B-cell lymphoma affecting predominantly children and young adults. Blood 2011;118(1):139-47.
3. Vasmatzis G, Johnson SH, Knudson RA, et al. Genome-wide analysis reveals recurrent structural abnormalities of TP63 and other p53-related genes in peripheral T-cell lymphomas. Blood 2012;120(11):2280-2289.
4. Crescenzo R, Abate F, Lasorsa E, et al. Convergent Mutations and Kinase Fusions Lead to Oncogenic STAT3 Activation in Anaplastic Large Cell Lymphoma. Cancer Cell 2015;27(4):516-532.
5. Hu G, Dasari S, Asmann YW, et al. Targetable fusions of the FRK tyrosine kinase in ALK-negative anaplastic large cell lymphoma. Leukemia 2018;32(2):565-569.

**Table S2: All mutations identified by WES in patient 1 (**Attached as a separate excel file).

Only variants called across all 4 variant callers were considered. Variants are only reported if abiding to the following criteria: (i) covered by >20 reads, (ii) predicted consequence is not synonymous or within an intron or a noncoding transcript, (iii) impact predicted moderate or high, (iv) present in the vep gnomAD database at a frequency <0.01 and (v) present in the gnomAD online browser (version 4.1.0) at a frequency <0.0001. All variants were manually validated in IGV (version 2.19.1). The annotations from all variant callers (Mutect2, Lofreq, Octopus and VarScan2) are listed in this table.
